# Long-term Intake of Statins or Renin-Angiotensin-System Inhibitors and Their Association With Parameters of Sarcopenia in Older Persons

**DOI:** 10.1101/2025.08.01.25332603

**Authors:** Tansu M. I. Aydoğan, Sarah Toepfer, Dominik Spira, Reinhold Kreutz, Ilja Demuth

## Abstract

**Background:** The impact of common cardiovascular medications on sarcopenia in older persons is unclear. The aim of this population-based cohort study was to investigate the relationship between long-term intake of statins and renin-angiotensin-system inhibitors (RASi), i.e. angiotensin-converting enzyme inhibitors (ACEi) or angiotensin receptor blockers (ARB), and established muscle mass and function parameters.

**Methods:** Community dwelling older adults (n=1,083, 52% women, 68.3 ± 3.5 years at baseline) from the Berlin Aging Study II (BASE-II) with a mean follow-up of 7.4 ± 1.5 years were analyzed. Users of statins or RASi were compared to non-users. Appendicular lean mass (ALM) was measured via dual-energy X-ray absorptiometry and related to body mass index (ALM/BMI) and height^2^ (SMI). Hand grip strength (HGS) was assessed and related to arm muscle mass. The Timed “Up and Go” and Tinetti mobility test were used to evaluate physical performance. Adjusted linear regressions of drug use on muscle mass, strength, and function were conducted.

**Results:** In linear regression analysis, statin use was neither associated with any of the muscle mass nor with function parameters. In contrast, RASi users had significantly lower ALM/BMI (β= −0.036, p=0.004). When stratified by sex, the use of RASi was significantly associated with lower ALM/BMI in women (β= −0.033, p=0.029) but not in men (β= −0.037, p=0.072).

**Conclusion:** Although statins have been associated with adverse muscle events, their use in older adults was not significantly associated with muscle mass or function parameters. In contrast, use of RASi was associated with a lower muscle mass phenotype.

**Graphical Abstract:** 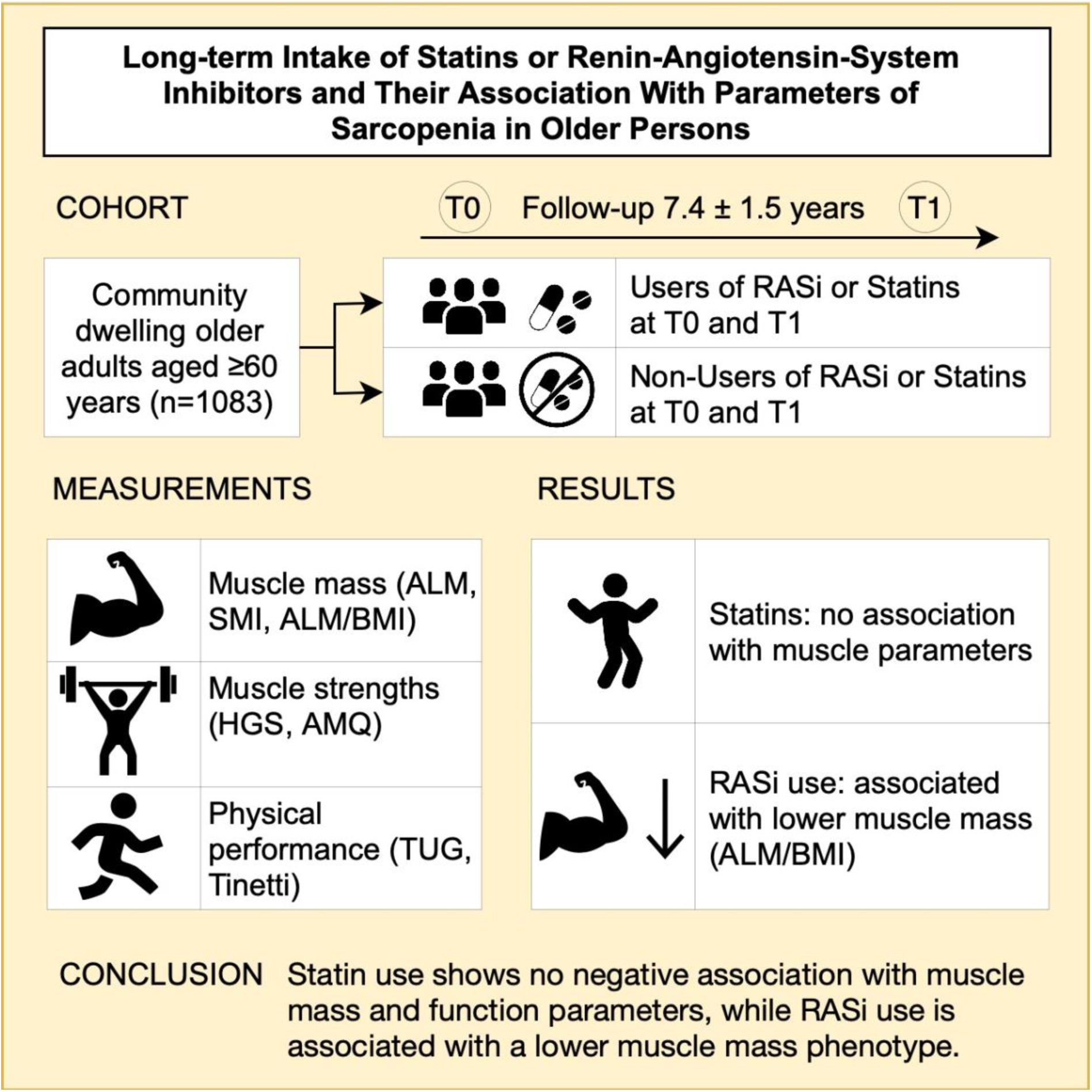

## Introduction

Sarcopenia is a skeletal muscle disorder, characterized by loss of muscle mass, strength and function, often occurring at an older age.^1–4^ It is associated with a higher risk of falls, limited physical abilities and mortality,^1,5^ contributing to a reduced quality of life and higher healthcare burden.^1,6^

To date, the most effective strategy to prevent and treat sarcopenia is resistance training, while no drug has yet been explicitly approved for this indication.^7–9^ Moreover, resistance training is also recommended in recent hypertension guidelines for blood pressure lowering.^10,11^ However, limited accessibility and motivation can be barriers for implementing resistance training in routine care, therefore pharmacotherapy is being explored as a possible alternative.^9,12^ A recent meta-analysis revealed that in older people sarcopenia correlates with hypertension.^13^ Investigating the potential impact of medications including common drugs to treat or prevent cardiovascular disease (CVD) such as statins and inhibitors of the renin-angiotensin system (RASi), i.e. angiotensin-converting enzyme inhibitors (ACEi) and angiotensin receptor blockers (ARB) on muscle parameters is of clinical interest since both beneficial and detrimental effects could affect treatment decisions.^9^

Statins are associated with muscle side effects including myopathy and myalgia^14,15^ but studies on their relationship with sarcopenia yielded contradictory results, ranging from detrimental^16^ to beneficial effects,^17,18^ whereas some longitudinal studies^19,20^ and randomized controlled trials (RCTs)^21,22^ showed no association between statin use and muscle mass or functional parameters.

Regarding ACEi, our group previously conducted a cross-sectional analysis using data of 838 participants from the Berlin Aging Study II (BASE-II), and found no association between intake of ACEi and muscle mass, strength and function, after adjusting for age, fat mass, hypertension, and morbidities.^23^

Other studies reported conflicting results for ACEi and ARB, with some suggesting favorable associations with muscle mass, strength or function,^24–27^ and others including RCTs and meta-analyses^7,28,29^ showing no positive or even a negative association.

Given these contradictory findings, real world data may complement RCT results due to larger and more diverse cohorts and longer follow-up periods.^30^ We aimed to investigate whether long-term intake of statins or RASi is associated with muscle mass, strength and physical performance in community-dwelling older adults of BASE-II. Additionally, potential sex differences were examined.

## Methods

### Data Availability

Due to concerns for participant privacy, data are available only upon reasonable request. Please contact Ludmila Müller, scientific coordinator, at, for additional information.

### Participants

Participants of the BASE-II were originally recruited from the greater Berlin metropolitan area.^31^ 1083 participants were included in this longitudinal study using baseline (T0) and follow-up (T1) data of BASE-II. The follow-up assessment on average 7.4 years after baseline was part of the GendAge study.^32^ Further details about both studies have been published elsewhere.^31–33^ All participants gave written informed consent. The Ethics Committee of Charité – Universitätsmedizin Berlin approved the study (approval numbers EA2/029/09 and EA2/144/16). The study was conducted in accordance with the Declaration of Helsinki and was registered in the German Clinical Trials Registry as DRKS00009277 and DRKS00016157.

### Body composition

Weight and height were examined using a measuring station and used to calculate the body mass index (BMI) [kg/m^2^]. Dual-energy X-ray absorptiometry (DXA) was used to obtain the appendicular lean mass (ALM) [kg].^23,34^ The skeletal muscle mass index (SMI = ALM/height^2^) [kg/m^2^], and the BMI adjusted parameter ALM/BMI^1^ were calculated. Detailed information and cut-off values can be obtained from the supplementary methods.^1,35,36^

### Physical performance

The Tinetti mobility test (Tinetti) and timed “Up and Go” test (TUG) [s] were performed as functional parameters.^37^ The hand grip strength (HGS) [kg]^38^ was assessed and arm muscle quality (AMQ) was derived as the ratio of HGS and arm lean mass.^39^ Further information can be obtained from the supplementary methods (Table S1).

### Questionnaires about physical performance and lifestyle

Participants self-evaluated whether they were rarely or never physically active (physical inactivity). Regular alcohol consumption (yes or no) and smoking considered as pack-years (number of cigarette packs smoked per day times years smoked) and current smoking (yes or no)^40^ were recorded.

### Medication intake grouping

The intake of statins, ARB, and ACEi was extracted at T0 and T1. Users took the drug at T0 and T1, whereas non-users neither took it at T0 nor at T1 (Panel A of Figure 1). The intake between T0 and T1 was assumed to be continuous. Participants taking the drug only at one time point were excluded. Users of either an ACEi or ARB were combined into one group, i.e. renin-angiotensin-system inhibitor group (RASi). Further information can be obtained from the supplementary methods. Polypharmacy was defined as the regular use of at least five drugs.^41^

**Figure 1.**
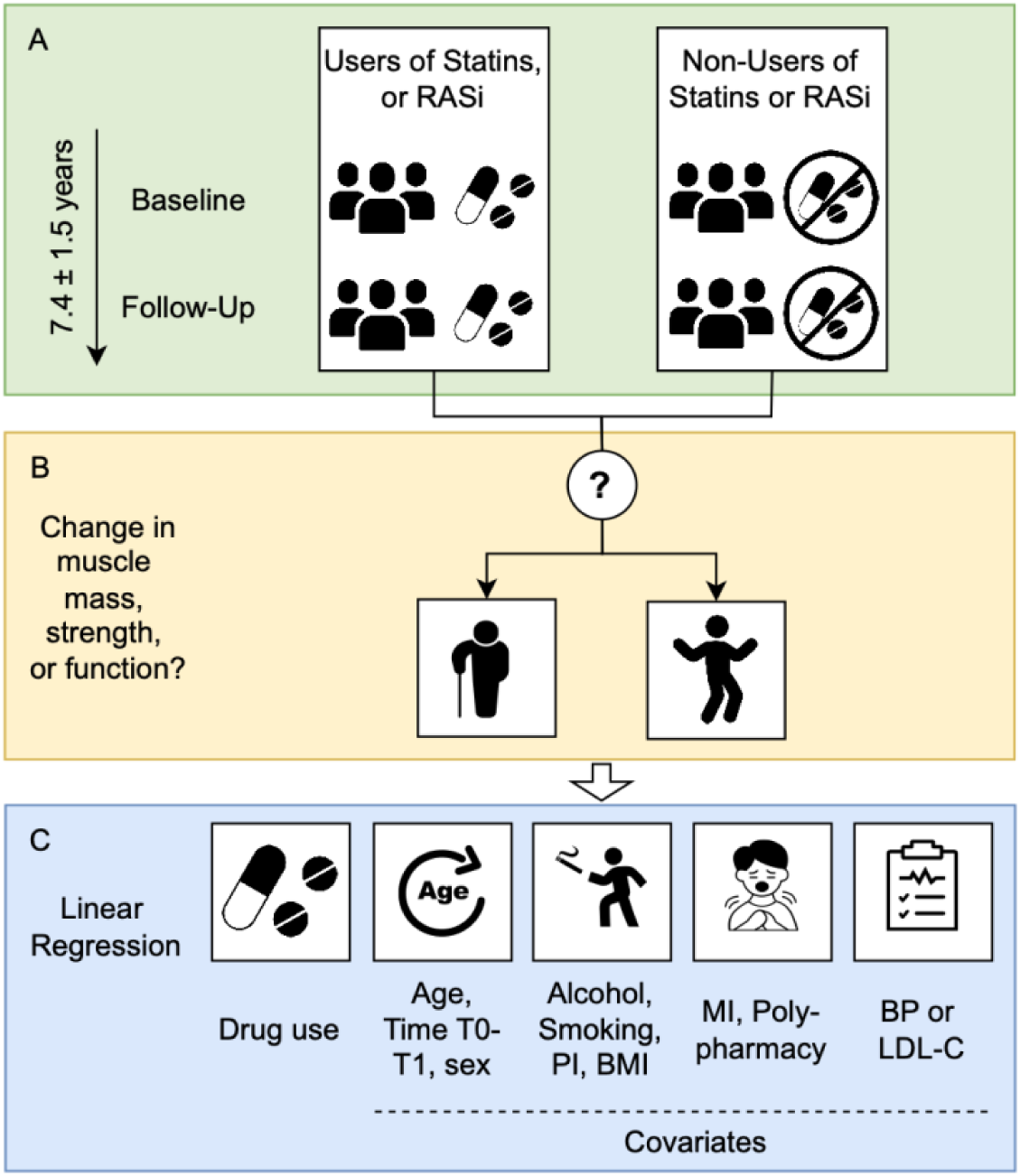
Workflow. A) Participant grouping according to drug intake. B) Outcomes of interest. C) Linear regression of drug use on muscle mass, strength and functional parameters with 5 steps of adding covariates. RASi: renin-angiotensin-system inhibitor, T0: baseline, T1: follow-up, Alcohol: regular alcohol consumption, Smoking: pack-years, PI: physical inactivity, BMI: body mass index, MI: morbidity index, BP: blood pressure, LDL-C: low density lipoprotein cholesterol

### Morbidities

The morbidity index (MI) is an adjusted version of the Charlson comorbidity index.^42,43^ CVD was defined as at least one of myocardial infarction, stroke, peripheral arterial disease or coronary heart disease at baseline. Furthermore, hypertension, type II diabetes, and heart failure at T0 were assessed. Hypertension was diagnosed by use of antihypertensive medication or systolic blood pressure (BP) ≥140 mmHg and/or diastolic BP ≥90 mmHg, as described before.^44^ Diagnosis criteria of type II diabetes mellitus^45^ are described in the supplementary methods.

### Laboratory parameters

Thyroid stimulating hormone (TSH), C-reactive protein (CRP),^43^ and low-density lipoprotein cholesterol (LDL*-*C) were determined in a certified laboratory. Systolic and diastolic BP were measured with an automatic upper arm BP monitor. One measurement was taken on each arm while sitting, and the mean was calculated between both arms. The estimated glomerular filtration rate (eGFR) was calculated using serum creatinine according to the Chronic Kidney Disease Epidemiology Collaboration (CKD-EPI) equation.^46^

### Statistics

Statistical analyses were performed using IBM SPSS Statistics, version 29 (IBM Corp., Armonk, N.Y., USA). Since muscle mass, strength, and physical performance are subject to sex differences, the analyses were additionally stratified by sex.^1,3,5,47^ The t-test, Welch’s test, Mann-Whitney U test, or Chi^2^-test were conducted to evaluate differences between groups (women and men or drug users and non-users).

To investigate whether drug intake (statins or RASi) is associated with ALM, SMI, ALM/BMI, HGS, AMQ, TUG time and Tinetti score, linear regressions were conducted for each outcome with 5 steps, to increasingly adjust for the potential confounders age, individual time between T0 and T1, sex, regular alcohol consumption, pack-years, physical inactivity, BMI, MI, polypharmacy, and lastly systolic BP for RASi and LDL-C for statins (Panel C of Figure 1). These regression analyses were conducted for the overall population and stratified by sex. For any significant association of RASi intake with an outcome parameter, separate analysis for users of either ACEi or ARB was conducted. Besides this, TSH was added to the 5^th^ model for sensitivity analysis of functional parameters (HGS, AMQ, TUG, Tinetti test). Predictor variables were used from T0, criterion variables from T1. Participants with missing data were excluded from regression analyses. Statistical significance was set a priori at p <0.05, with no adjustment for multiple testing due to the exploratory nature of this study. This study was reported in accordance with the STROBE guidelines.

For outcomes which were significantly associated with drug intake in regression analyses, sex-stratified box-whisker plots were generated in R version 4.3.1^48^ using RStudio version 2023.06.1.524.^49^

## Results

### Population characteristics

1,083 participants (52% women) with a mean age of 68.3 ± 3.5 years at T0 were reassessed on average 7.4 ± 1.5 years (range 3.9 to 10.4 years) later (T1). Table 1 shows the characteristics of the overall study population and stratified by sex. Hypertension at baseline was prevalent in 72.8% of participants and significantly more frequent in men than in women. Type II Diabetes and CVD were also significantly more prevalent in men, whereas heart failure was more frequent in women. Among those with assumed continuous drug intake between T0 and T1 (“users”), 101 participants were in the statins group, and 253 in the RASi group. As expected, men had significantly higher values in muscle mass and strength parameters, namely ALM/BMI (Table 1), ALM, SMI and HGS (Table S2). DXA data from both time points were available for 652 participants (n=350 women). Further details on the distribution of participants by drug use and availability of DXA data are shown in Figure 2.

**Figure 2.**
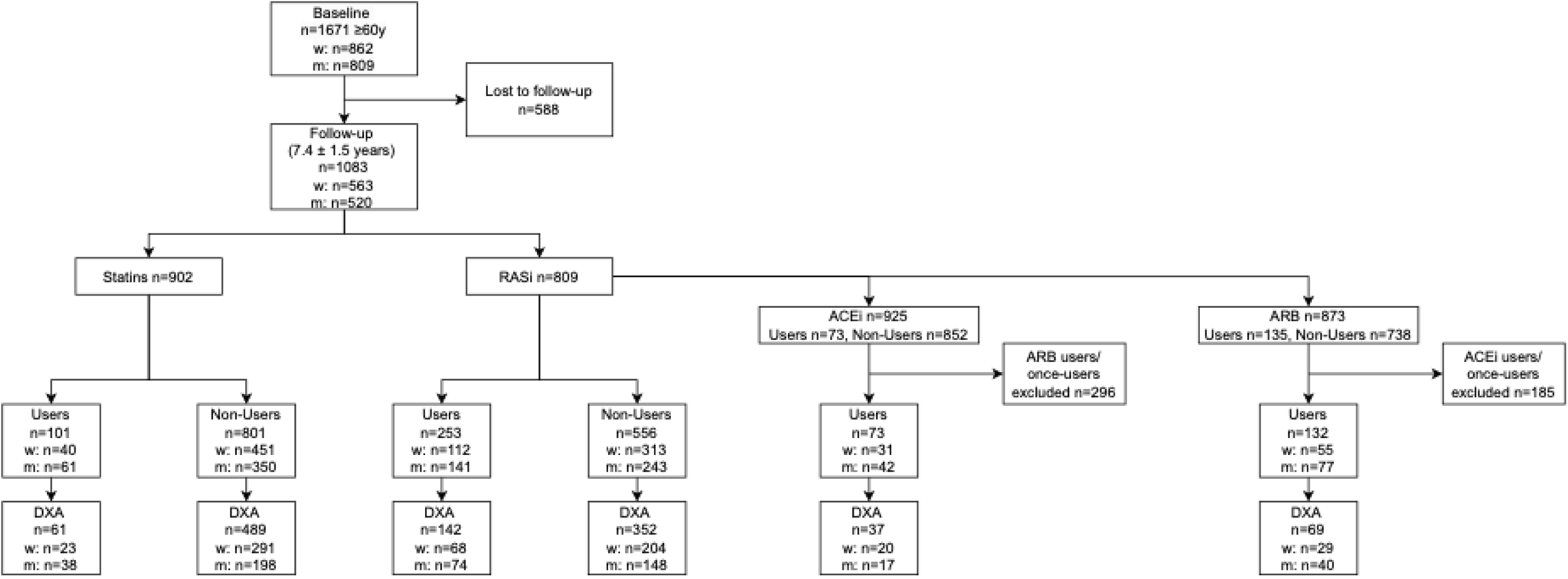
Flow-chart showing the selection of participants and the distribution between groups of drug users and non-users, the number of participants with DXA data available, and gender distribution. w: women, m: men, ACEi: angiotensin-converting enzyme inhibitor, ARB: angiotensin receptor blocker, RASi: renin-angiotensin-system inhibitor, DXA: dual-energy X-ray absorptiometry

**Table 1.**
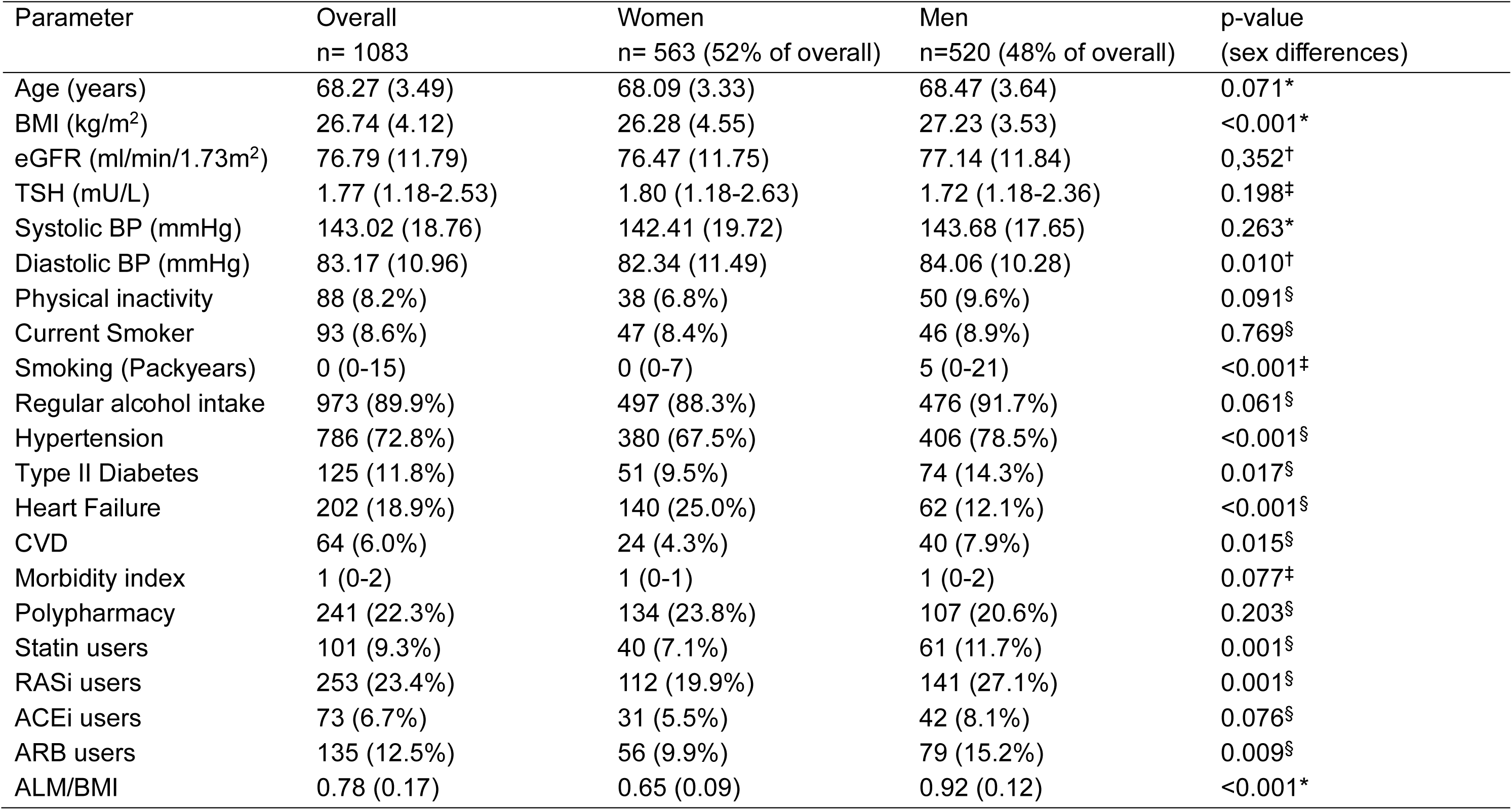
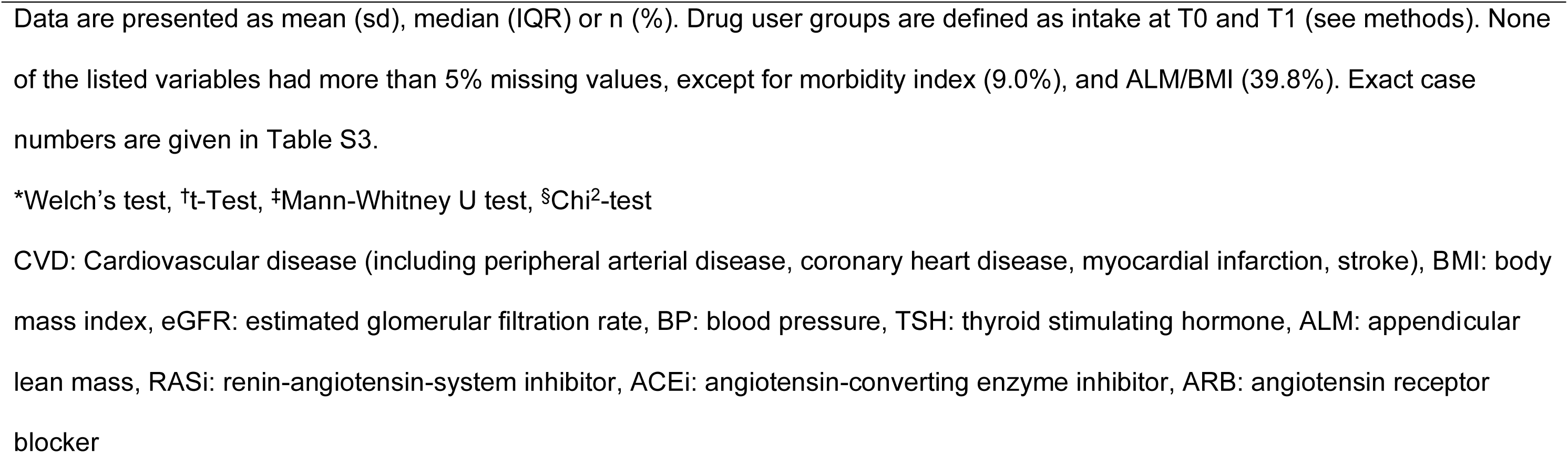
Characteristics of the overall study population and according to sex.

| Parameter | Overall<br>n= 1083 | Women<br>n= 563 (52% of overall) | Men<br>n=520 (48% of overall) | p-value<br>(sex differences) |
| --- | --- | --- | --- | --- |
| Age (years) | 68.27 (3.49) | 68.09 (3.33) | 68.47 (3.64) | 0.071* |
| BMI (kg/m<sup>2</sup>) | 26.74 (4.12) | 26.28 (4.55) | 27.23 (3.53) | <0.001* |
| eGFR (ml/min/1.73m<sup>2</sup>) | 76.79 (11.79) | 76.47 (11.75) | 77.14 (11.84) | 0.352<sup>†</sup> |
| TSH (mU/L) | 1.77 (1.18-2.53) | 1.80 (1.18-2.63) | 1.72 (1.18-2.36) | 0.198<sup>‡</sup> |
| Systolic BP (mmHg) | 143.02 (18.76) | 142.41 (19.72) | 143.68 (17.65) | 0.263* |
| Diastolic BP (mmHg) | 83.17 (10.96) | 82.34 (11.49) | 84.06 (10.28) | 0.010<sup>†</sup> |
| Physical inactivity | 88 (8.2%) | 38 (6.8%) | 50 (9.6%) | 0.091<sup>§</sup> |
| Current Smoker | 93 (8.6%) | 47 (8.4%) | 46 (8.9%) | 0.769<sup>§</sup> |
| Smoking (Packyears) | 0 (0-15) | 0 (0-7) | 5 (0-21) | <0.001<sup>‡</sup> |
| Regular alcohol intake | 973 (89.9%) | 497 (88.3%) | 476 (91.7%) | 0.061<sup>§</sup> |
| Hypertension | 786 (72.8%) | 380 (67.5%) | 406 (78.5%) | <0.001<sup>§</sup> |
| Type II Diabetes | 125 (11.8%) | 51 (9.5%) | 74 (14.3%) | 0.017<sup>§</sup> |
| Heart Failure | 202 (18.9%) | 140 (25.0%) | 62 (12.1%) | <0.001<sup>§</sup> |
| CVD | 64 (6.0%) | 24 (4.3%) | 40 (7.9%) | 0.015<sup>§</sup> |
| Morbidity index | 1 (0-2) | 1 (0-1) | 1 (0-2) | 0.077<sup>‡</sup> |
| Polypharmacy | 241 (22.3%) | 134 (23.8%) | 107 (20.6%) | 0.203<sup>§</sup> |
| Statin users | 101 (9.3%) | 40 (7.1%) | 61 (11.7%) | 0.001<sup>§</sup> |
| RASi users | 253 (23.4%) | 112 (19.9%) | 141 (27.1%) | 0.001<sup>§</sup> |
| ACEi users | 73 (6.7%) | 31 (5.5%) | 42 (8.1%) | 0.076<sup>§</sup> |
| ARB users | 135 (12.5%) | 56 (9.9%) | 79 (15.2%) | 0.009<sup>§</sup> |
| ALM/BMI | 0.78 (0.17) | 0.65 (0.09) | 0.92 (0.12) | <0.001* |

Data are presented as mean (sd), median (IQR) or n (%). Drug user groups are defined as intake at T0 and T1 (see methods). None of the listed variables had more than 5% missing values, except for morbidity index (9.0%), and ALM/BMI (39.8%). Exact case numbers are given in Table S3.
\*Welch's test, <sup>†</sup>t-Test, <sup>‡</sup>Mann-Whitney U test, <sup>§</sup>Chi<sup>2</sup>-test
CVD: Cardiovascular disease (including peripheral arterial disease, coronary heart disease, myocardial infarction, stroke), BMI: body mass index, eGFR: estimated glomerular filtration rate, BP: blood pressure, TSH: thyroid stimulating hormone, ALM: appendicular lean mass, RASi: renin-angiotensin-system inhibitor, ACEi: angiotensin-converting enzyme inhibitor, ARB: angiotensin receptor blocker

### Statin group

Regression analyses revealed no significant associations between statin use and muscle mass, strength or functional outcome parameters in the fully adjusted regression models (Table S4). This remained unchanged after adding TSH as an additional covariable (Table S4) or stratifying by sex (Table S5).

### RASi group

In the fully adjusted regression model 5, RASi intake was significantly associated with lower ALM/BMI (β= −0.036, p=0.004) (Table 2). There was no association between RASi intake and the remaining parameters of muscle mass, strength or function, which did not change when TSH was included (Table S4).

**Table 2.** Linear Regression Analyses of ALM/BMI on RASi intake in the overall population and stratified by sex.

| | n | $\beta$ | 95% CI LL | 95% CI UL | p |
| --- | --- | --- | --- | --- | --- |
| Overall | 431 | -0.036 | -0.060 | -0.012 | 0.004 |
| Women | 233 | -0.033 | -0.063 | -0.003 | 0.029 |
| Men | 198 | -0.037 | -0.078 | 0.003 | 0.072 |

Analyses were adjusted for age, age difference T0-T1, sex (overall analysis), current alcohol consumption, pack-years, physical inactivity, morbidity index, polypharmacy, and blood pressure.
CI: confidence interval, LL: lower limit, UL: upper limit, RASi: renin-angiotensin-system inhibitor, ALM/BMI: appendicular lean mass divided by body mass index

Stratification of RASi users by sex is visualized in Figure 3, showing individual ALM/BMI values. As expected, ALM/BMI values are lower in women than in men. There are significant differences between RASi users and non-users in both genders (Figure 3), but after adjustment, regression analyses revealed a significant association between RASi use and lower ALM/BMI in women, but not in men (Table 2). Adding TSH in sensitivity analysis, did not reveal additional associations (Table S5). All other results of the regression analyses (model 5) were not significant and are provided in Table S4 (overall) and Table S5 (sex-stratified).

**Figure 3.**
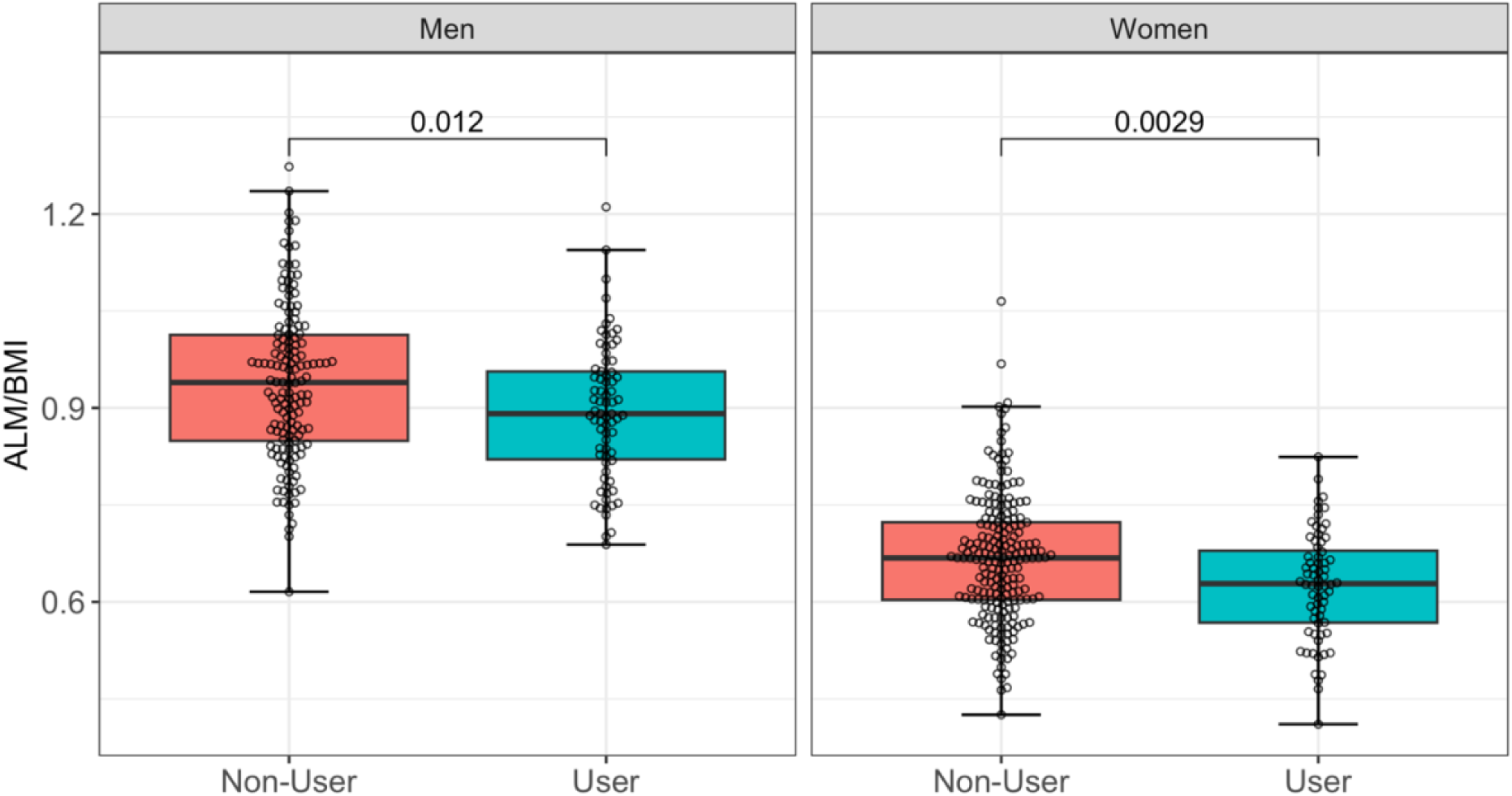
Box plots with distribution of participants showing differences in ALM/BMI according to the use of RASi, stratified by sex. Differences between users and non-users were tested by Mann-Whitney U test. Plots are only shown for ALM/BMI, since this parameter shows significant results in the regression analyses. Men: Non-User n= 148, User n=74; Women: Non-User n=204, User n=68. RASi: renin-angiotensin-system inhibitor, ALM/BMI: appendicular lean mass divided by body mass index

When ACEi and ARB were analyzed separately in the overall study population, ACEi use was associated with lower ALM/BMI (β= −0.046, p=0.027), while ARB use showed no associations (Table S4). Sex-stratified analyses for ACEi and ARB subgroups were not feasible due to insufficient sample sizes.

## Discussion

In this study, we assessed whether the long-term intake of statins or RASi, as indicated by reporting use of these medications at both baseline and during a mean follow-up of 7.4 ± 1.5 years, is associated with sarcopenia parameters in a sample of older people with a mean age of 68.3 ± 3.5 years and a high prevalence of arterial hypertension. While statin use was not associated with muscle mass, strength or physical performance parameters, we found RASi use to be significantly associated with lower ALM/BMI (β= −0.036, p=0.004) in the overall study population. When stratified by sex, RASi use was significantly associated with lower ALM/BMI in women (β= −0.033, p=0.029) but not in men (Table 2). In the overall population, ACEi use was associated with lower ALM/BMI (β= −0.046, p=0.027), while ARB showed no associations (Table S4).

The absence of an association between long-term-statin use and muscle outcomes in our study including older people is reassuring, since statin use has been associated with adverse muscular effects such as pain and cramps, as well as more severe conditions such as myopathy and rhabdomyolysis, which can lead to intolerance and non-adherence, despite CV risks.^14,15,50^ Interestingly, recent meta-analyses on the nocebo effect of statin therapy have shown that muscle symptoms due to statin therapy are often overestimated.^14,50^ Therefore, given the rising use of statins in older adults^51^ at high cardiovascular risk as recommended in American and European guidelines,^10,11,52^ it is encouraging that our findings suggest that statin use is not a risk factor for decline in muscle mass, strength or function which may progress subclinically compared to more severe side effects such as myopathy.

Our findings align with the Hertfordshire Cohort Study,^20^ and the NILS-LSA,^19^ which also investigated community-dwelling older adults. The contradictory results of other studies may be due to heterogeneity in populations, underlying diseases or interventions, limiting comparability.^17,18,21,22^

Regarding the antihypertensive medications examined in this study, intake of RASi showed a significant association with lower ALM/BMI overall and in women (Table 2). The lack of significance in men may reflect the smaller sample size, while the beta-value points in the same direction as for women. Lower mean ALM/BMI in RASi users of both genders compared to non-users was moreover indicated in Figure 3. Two studies found beneficial effects of both ACEi and ARB intake in women^25^ or both genders.^53^ Generally, evidence on sex-specific differences in the effects of RASi is limited,^54^ particularly regarding muscle parameters. Possible explanations include hormonal influences on the renin-angiotensin system or sex-specific differences in pharmacokinetics and pharmacodynamics.^54,55^

When considering RASi subgroups, the use of ACEi, but not ARB, showed significantly lower ALM/BMI in the overall study population despite the smaller group size (Table S4). In our previous cross-sectional analysis of BASE-II, ACEi use was not associated with ALM/BMI after adjustment for potential confounders^23^ suggesting that effects on muscle mass (ALM/BMI) might only become apparent after many years of regular ACEi intake. One cross-sectional study found no association between leg muscle mass and ACEi.^24^ Other studies investigating ACEi or ARB focused on functional tests^20,25,26,28,29^ rather than muscle mass. However, the negative β-value in the ARB regression analysis may indicate a trend toward lower ALM/BMI in long-term ARB users as well (Table S4).

Alongside ALM/BMI, we assessed SMI (ALM adjusted for height^2^) and found no significant differences between users and non-users of anti-hypertensive drugs. This highlights the debate over optimal muscle metrics for sarcopenia.^1–5,8^ However, our previous study showed that ALM/BMI was associated with physical limitations and frailty, whereas SMI was not,^56^ suggesting that ALM/BMI is better suited to reflect sarcopenia.

According to consensus statements, physical performance should be evaluated in addition to muscle mass.^1–5,8^ While some studies found beneficial associations between RASi use and physical performance,^53,57^ others including meta-analysis of RCTs did not find associations,^29^ which aligns with our results.

This study is limited by several factors which we would like to acknowledge. Firstly, the study relies on the assumption of continuous medication intake if documented at both assessment time points. Therefore, continuous intake it is not guaranteed, but in our view very likely. Secondly, selection bias may have occurred due to participant limitations for DXA measurements (e.g. implants), exclusion due to drug intake at only one point of time, and loss to follow-up, all of which reduced statistical power. However, no significant difference in overall morbidity at baseline was found between participants lost to follow-up and those reassessed.^45^ Besides, analyses with less healthy participants might have yielded more pronounced results than our community-dwelling cohort, which was over-average healthy,^58^ potentially limiting the identification of impairments.^23^ The heterogeneity in health status and the use of other medications was addressed by adjusting for morbidity index and polypharmacy. Thirdly, it is being discussed whether other methods for muscle mass measurement are better suited to identify sarcopenia than DXA.^4,5^ Finally, as an exploratory study, multiple testing was not corrected for, requiring confirmatory research.

Strengths of the study include the use of comprehensive longitudinal data with a follow-up of 7.4 ± 1.5 years, which is longer than in most previous studies on the drugs investigated and sarcopenia, as well as its relatively large, balanced sample which enabled sex-stratified analyses. Both, a large sample size and long follow-up are better feasible in observational studies like ours than in RCTs.^30^

## Perspectives

Statin use in older people was not associated with indicators of sarcopenia, which is reassuring considering its muscle side effects. RASi intake, on the other hand, was associated with lower muscle mass in our study. Future confirmatory research is needed and should focus on longer follow-up periods, and balanced participation of women and men. Due to the high prevalence of CVD in older people, understanding the effects of commonly used drugs on sarcopenia is clinically relevant. This also calls for an internationally standardized definition and validated diagnostic parameters for sarcopenia.

## Novelty and Relevance

### What Is New?

- Long-term treatment with renin-angiotensin-system inhibitors (RASi) was associated with lower muscle mass in older persons.
- Statin treatment showed no association with muscle mass, strength or function.
- This study had a longer follow-up than most studies on this topic.

### What is relevant?

- Hypertensive patients are at risk of sarcopenia.
- Long-term treatment with RASi may reduce muscle mass in older adults.

### Clinical Implications

- If confirmed, the possible effects of hypertension treatment on sarcopenia should be considered in clinical decision-making.

## Acknowledgements

The authors are grateful to all participants of BASE-II.

## Sources of Funding

This article uses data from the BASE-II. BASE-II was supported by the German Federal Ministry of Education and Research under grant numbers #01UW0808; #16SV5536K, #16SV5537, #16SV5538, #16SV5837, #01GL1716A, and #01GL1716B.

## Disclosures

RK reports received honoraria for consultancy and lectures from AstraZeneca, Bayer, Merck, Krka, Menarini Group, ProMed, PolPharma, Recor, Servier, and Zentiva. The other authors declare no conflicts of interest.

## Author contributions

Conceptualization: T.M.I.A., D.S., R.K., I.D.; Data curation: T.M.I.A., S.T., D.S., I.D.; Formal analysis: T.M.I.A.; Funding acquisition: I.D.; Resources: I.D.; Supervision: D.S., R.K., I.D.; Visualization: T.M.I.A.; Writing first draft: T.M.I.A.; Reviewed and approved the manuscript: all authors.

## Supplemental Material

Supplemental Methods

Tables S1-S5

Reference 59

## Non-standard Abbreviations and Acronyms

ACEi: angiotensin-converting enzyme inhibitor
ALM: appendicular lean mass
ALM/BMI: appendicular lean mass divided by body mass index
AMQ: arm muscle quality
ARB: angiotensin receptor blocker
BASE-II: Berlin Aging Study II
DXA: dual-energy X-ray absorptiometry
HGS: hand grip strength
RASi: renin-angiotensin-system inhibitor
SMI: skeletal muscle mass index
T0: baseline measurement time point
T1: follow-up measurement time point
TUG: timed “Up and Go” test

## Notes

### Clinical Trial

The study was registered in the German Clinical Trials Registry as DRKS00009277 and DRKS00016157.

### Funding Statement

This article uses data from the Berlin Aging Study II (BASE-II). BASE-II was supported by the German Federal Ministry of Education and Research under grant numbers #01UW0808; #16SV5536K, #16SV5537, #16SV5538, #16SV5837, #01GL1716A, and #01GL1716B.

### Author Declarations

The Ethics Committee of Charité - Universitätsmedizin Berlin approved the study (approval numbers EA2/029/09 and EA2/144/16). The study was conducted in accordance with the Declaration of Helsinki and was registered in the German Clinical Trials Registry as DRKS00009277 and DRKS00016157.

